# Influence of vitamin D supplementation on growth, body composition, pubertal development and spirometry in South African schoolchildren: a randomised controlled trial (ViDiKids)

**DOI:** 10.1101/2023.11.29.23299226

**Authors:** Keren Middelkoop, Lisa K Micklesfield, Justine Stewart, Neil Walker, David A Jolliffe, Amy E Mendham, Anna K Coussens, James Nuttall, Jonathan C Y Tang, William D Fraser, Waheedullah Momand, Cyrus Cooper, Nicholas C Harvey, Robert J Wilkinson, Linda-Gail Bekker, Adrian R Martineau

**Affiliations:** Desmond Tutu HIV Centre, Institute of Infectious Disease & Molecular Medicine, University of Cape Town, Observatory, South Africa; Department of Medicine, University of Cape Town, Observatory, South Africa; Health through Physical Activity, Lifestyle and Sport Research Centre (HPALS), Division of Physiological Sciences, Department of Human Biology, Faculty of Health Sciences, University of Cape Town, Cape Town, South Africa; SAMRC/Wits Developmental Pathways for Health Research Unit, Department of Paediatrics, Faculty of Health Sciences, University of the Witwatersrand, Johannesburg, South Africa; Wolfson Institute of Population Health, Barts and The London School of Medicine and Dentistry, Queen Mary University of London, London, UK; Blizard Institute, Barts and The London School of Medicine and Dentistry, Queen Mary University of London, London, UK; Centre for Infectious Diseases Research in Africa and Institute of Infectious Disease and Molecular Medicine, Faculty of Health Sciences, University of Cape Town, Observatory 7925, South Africa; Infectious Diseases and Immune Defence Division, Walter and Eliza Hall Institute of Medical Research, Parkville, Australia; Paediatric Infectious Diseases Unit, Red Cross War Memorial Children’s Hospital and the Department of Paediatrics and Child Health, University of Cape Town, Cape Town, South Africa; Norwich Medical School, University of East Anglia, Norwich Research Park, Norwich, UK; Departments of Laboratory Medicine, Clinical Biochemistry and Departments of Diabetes and Endocrinology, Norfolk and Norwich University Hospital NHS Foundation Trust, Norwich, UK; MRC Lifecourse Epidemiology Centre, University of Southampton, Southampton UK; NIHR Southampton Biomedical Research Centre, University of Southampton, Southampton UK and University Hospital Southampton NHS Foundation Trust, Southampton, UK; The Francis Crick Institute, London, NW1 1AT, UK; Department of Infectious Diseases, Imperial College London, London, W12 0NN, UK

**Author notes:** To whom correspondence should be addressed at the Blizard Institute, Barts and The London School of Medicine and Dentistry, Queen Mary University of London, 4 Newark St, London E1 2AT, UK. These authors contributed equally (Keren Middelkoop, Lisa K Micklesfield).

## Abstract

**Objective:** To determine whether weekly oral vitamin D supplementation influences growth, body composition, pubertal development or spirometric outcomes in South African schoolchildren.

**Design:** Phase 3 double-blind randomised placebo-controlled trial (clinicaltrials.gov registration no. NCT02880982).

**Setting:** Socio-economically disadvantaged peri-urban district of Cape Town, South Africa

**Participants:** 1682 children of Black African ancestry attending government primary schools and aged 6-11 years at baseline.

**Interventions:** Oral vitamin D_3_ (10,000 IU/week) vs. placebo for 3 years

**Main outcome measures:** height-for-age and body mass index-for-age, measured in all participants); Tanner scores for pubertal development, spirometric lung volumes and body composition, measured in a subset of 450 children who additionally took part in a nested sub-study.

**Results:** Mean serum 25-hydroxyvitamin D_3_ concentration at 3-year follow-up was higher among children randomised to receive vitamin D vs. placebo (104.3 vs. 64.7 nmol/L, respectively; mean difference [MD] 39.7 nmol/L, 95% CI 37.6 to 41.9 nmol/L). No statistically significant differences in height-for-age z-score (adjusted MD [aMD] −0.08, 95% CI −0.19 to 0.03) or body mass index-for-age z-score (aMD −0.04, 95% CI −0.16 to 0.07) were seen between vitamin D vs. placebo groups at follow-up. Among sub-study participants, allocation to vitamin D vs. placebo did not influence pubertal development scores, % predicted forced expiratory volume in 1 second (FEV1), % predicted forced vital capacity (FVC), % predicted FEV1/FVC, fat mass or fat-free mass.

**Conclusions:** Weekly oral administration of 10,000 IU vitamin D_3_ boosted vitamin D status but did not influence growth, body composition, pubertal development or spirometric outcomes in South African schoolchildren.

**KEY MESSAGES:** *What is already known on this topic?:*

- Observational studies have reported independent associations between vitamin D deficiency in childhood and slower linear growth, reduced lean mass, obesity and precocious puberty.
- A phase 2 clinical trial conducted in Mongolia reported that a 6-month course of vitamin D supplementation increased height gain in 113 vitamin D deficient schoolchildren aged 12-15 years; however, these results were not confirmed by a recent phase 3 trial conducted in the same setting.
- RCTs to determine effects of vitamin D supplementation on growth and development in schoolchildren have not been conducted in other settings.

*What this study adds:*

- This placebo-controlled phase 3 clinical trial, conducted in 1682 Black African schoolchildren in Cape Town, South Africa, showed that a 3-year course of weekly vitamin D supplementation was effective in elevating circulating 25-hydroxyvitamin D concentrations.
- However, this was not associated with any effect on linear growth, body composition, pubertal development or spirometric lung volumes.

*How this study might affect research, practice or policy:*

- Our findings do not support use of vitamin D supplementation as an intervention to influence child growth, body composition, pubertal development or spirometric lung volumes.

## INTRODUCTION

Middle childhood and early adolescence represent key periods for growth and development that has an important influence on stature and health outcomes in later adolescence and adulthood.^1^ Stunting in childhood has long been recognised to associate with multiple adverse long-term health outcomes,^2^ and is particularly common in lower-income countries.^3^ These settings have also witnessed an emerging epidemic of childhood obesity,^4^ which has in turn been associated with accelerated pubertal development.^5^ Interventions to alleviate these public health challenges are urgently needed.

Vitamin D is a fat-soluble micronutrient with pleotropic effects on human health.^6^ Observational studies have reported that vitamin D deficiency, as evidenced by low circulating concentrations of its major metabolite 25-hydroxyvitamin D (25[OH]D), associates with slower linear growth,^7^ reduced lean mass,^8^ childhood obesity^9–11^ and precocious puberty,^12,13^ potentially reflecting the ability of vitamin D to stimulate production of insulin-like growth factor 1^14^ and regulate adipogenesis.^15^ A phase 2 randomised controlled trial (RCT) conducted in Mongolia reported that a 6-month course of vitamin D supplementation increased height gain in 113 vitamin D deficient schoolchildren aged 12-15 years,^16^ but these results were not confirmed by a recent phase 3 RCT conducted in the same setting.^17^ RCTs to determine effects of vitamin D supplementation on growth and development in schoolchildren have not been conducted in other settings representing children at different risks of malnutrition, stunting and obesity. An opportunity to conduct such an investigation recently arose as part of the ViDiKids trial, a multicentre phase 3 RCT which investigated the effects of weekly oral administration of 10,000 IU vitamin D_3_ for 3 years on the primary outcome of tuberculosis infection in a cohort of 1682 schoolchildren aged 6-11 years living in a socio-economically disadvantaged peri-urban district of Cape Town, South Africa.^18^ Height-for-age z-scores and body mass index-for age z-scores were assessed in all participants (n=1682), and Tanner scores for pubertal development, spirometric lung volumes and body composition were assessed in a subset of 450 children who also took part in a nested sub-study.

## METHODS

### TRIAL DESIGN, SETTING, APPROVALS AND REGISTRATION

We conducted a multicentre phase 3 double-blind individually randomised placebo-controlled trial of weekly oral vitamin D supplementation in 23 government schools in Cape Town, South Africa, as previously described.^18,19^ The primary outcome was acquisition of tuberculosis infection, as evidenced by conversion of a QuantiFERON-TB Gold Plus (QFT-Plus) assay result from negative at baseline to positive at 3-year follow-up. The current manuscript reports effects of the intervention on pre-specified secondary outcomes relating to growth in all study participants, and body composition, pubertal development and spirometry in a subset of participants who additionally took part in a nested sub-study. The trial was sponsored by Queen Mary University of London, approved by the University of Cape Town Faculty of Health Sciences Human Research Ethics Committee (Ref: 796/2015) and the London School of Hygiene and Tropical Medicine Observational/Interventions Research Ethics Committee (Ref: 7450-2) and registered on the South African National Clinical Trials Register (DOH-27-0916-5527) and ClinicalTrials.gov (ref NCT02880982).

### PARTICIPANTS

Inclusion criteria for the main trial were enrolment in Grades 1-4 at a participating school; age 6 to 11 years at screening; and written informed assent / consent to participate in the main trial provided by children and their parent / legal guardian, respectively. Exclusion criteria for the main trial were a history of previous tuberculosis infection, active TB disease or any chronic illness other than asthma (including known or suspected HIV infection) prior to enrolment; use of any regular medication other than asthma medication; use of vitamin D supplements at a dose of more than 400 IU/day in the month before enrolment; plans to move away from study area within 3 years of enrolment; inability to swallow a placebo soft gel capsule with ease; and clinical evidence of rickets or a positive QFT-Plus assay result at screening. An additional inclusion criterion for the sub-study was enrolment in Grade 4 at a participating school.

### ENROLMENT

Full details of enrolment procedures are described in Supplementary Material. Eligible participants underwent measurement of weight using a digital floor scale (Charder Medical, Taichung City, Taiwan), height using a portable stadiometer (HM200P, Charder Medical) and waist circumference using a measuring tape. Sub-study participants also underwent spirometry according to ERS/ATS standards^20^ using a portable spirometer (Carefusion, San Diego, CA) and measurement of body composition by dual energy x-ray absorptiometry (DXA) as described below.

### RANDOMISATION AND BLINDING

Full details of randomisation and blinding procedures have been described previously, and are presented in Supplementary Material.^18,19^ Briefly, eligible and assenting children whose parents consented to their participation in the trial were individually randomised to receive a weekly capsule containing vitamin D_3_ or placebo for three years, with a one-to-one allocation ratio and randomisation stratified by school of attendance. Treatment allocation was concealed from participants, care providers and all trial staff (including senior investigators and those assessing outcomes) until completion of the trial to maintain the double-blind.

### INTERVENTION

Study medication comprised a 3-year course of weekly soft gel capsules manufactured by the Tishcon Corporation (Westbury, NY, USA), containing either 0.25 mg (10,000 international units) cholecalciferol (vitamin D_3_) in olive oil (intervention arm) or olive oil without any vitamin D_3_ content (placebo arm). Active and placebo capsules had identical appearance and taste. Capsules were taken under direct observation of study staff during school term time. Further details of administration of study medication are provided in Supplementary Material.

### FOLLOW-UP ASSESSMENTS

At 1-year, 2-year and 3-year follow-up, height, weight and waist circumference were measured as at baseline. At 3-year follow-up, sub-study participants were additionally invited to undergo repeat spirometry and DXA scanning, and to complete a Tanner self-assessment questionnaire for pubertal development.^21^

### OUTCOMES

The primary outcome for the main trial, reported elsewhere,^18^ was the QFT-Plus result at the end of the study. The following secondary outcomes were assessed for all participants: height-for-age, BMI-for-age, waist circumference-for-age and waist-to-height ratio (all participants). Additional outcomes assessed in sub-study participants only were: whole body fat mass, fat-free soft tissue mass, % predicted forced vital capacity (FVC), % predicted forced expiratory volume in one second (FEV1), % predicted FEV1/FVC, mean Tanner scores for pubic hair (males and females), external genitalia (males only) or breast development (females only), the proportion of participants reaching menarche by the end of the trial (females only) and mean age at menarche (females who reached menarche by the end of the trial only).

### DXA

Body composition (whole body fat mass and fat-free soft tissue mass) was assessed using a Hologic Discovery-W^®^ dual energy x-ray absorptiometry (DXA) scanner at the Sports Science Institute of South Africa, University of Cape Town. All scans were performed by a trained radiographer on one scanner (Hologic, Bedford, MA, USA) using standard procedures, and analysed using Apex software (Version 13.4.1). Quality assurance checks were carried out prior to scanning and generated coefficients of variation <0.5%.

### LABORATORY ASSESSMENTS

Serum concentrations of 25(OH)D_3_ were measured using liquid chromatography tandem mass spectrometry (LC-MS/MS) as previously described.^22^ Further details are presented in Supplementary Material.

### SAMPLE SIZE

Sample size for the main trial was predicated on power to detect an effect of the intervention on the primary outcome (the proportion of children with a positive QFT-Plus assay result at 3-year follow-up), as previously described.^18^ Sample size for the sub-study was predicated on power to detect an effect of the intervention on bone mineral content, as previously described.^19^

### STATISTICAL ANALYSES

Full details of statistical analyses are provided in Supplementary Material. Briefly, effects of treatment on age- and sex-adjusted z-scores for anthropometric outcomes were estimated by fitting allocation to vitamin D vs. placebo as the sole fixed effect in a mixed effects linear regression model with a random effect for repeated assessments of each individual participant and a random effect of school of attendance. Effects of treatment on DXA and spirometric outcomes were analysed using multi-level mixed models with adjustment for baseline values and a random effect for school. Tanner metrics were analysed in a similar fashion, but restricted to the applicable sex and without baseline adjustment. Pre-specified sub-group analyses were conducted to determine whether the effect of vitamin D supplementation was modified by sex (male vs. female), baseline deseasonalised 25(OH)D_3_ concentration (<75 vs. ≥75 nmol/L)^24^ and calcium intake (< vs. ≥ median value of 466 mg/day).^19^ An Independent Data Monitoring Committee reviewed accumulating serious adverse event data at 6-monthly intervals, and recommended continuation of the trial at each review. No interim efficacy analysis was performed.

## RESULTS

### PARTICIPANTS

A total of 2852 children were screened for eligibility from March 2017 to March 2019, of whom 2271 underwent QFT testing: 1682 (74.1%) QFT-negative children were randomly assigned to receive vitamin D_3_ (829 participants) or placebo (853 participants) as previously described.^18^ 450/1682 (26.8%) participants in the main trial also participated in the sub-study, of whom 228 vs. 222 participants were allocated to the vitamin D vs. placebo arms, respectively (Fig. 1). Table 1 presents baseline characteristics of children in the main trial and in the sub-study, overall and by study arm. Mean age was higher among participants in the sub-study vs. all those in the main trial (10.1 vs. 8.9 years, respectively), reflecting the fact that participation in the sub-study was restricted to children enrolled in Grade 4. Prevalence of obesity and stunting were lower in the sub-study vs. the main trial (3.8% vs 5.5%, and 6.4% vs. 11.1%, respectively. Baseline characteristics were otherwise well balanced for all participants in the main trial vs. those who additionally participated in the sub-study: 52.4% vs. 52.0% were female and mean serum 25(OH)D_3_ concentrations were 71.2 nmol/L vs. 70.0 nmol/L. Within the main trial and the sub-study, baseline characteristics of those randomised to vitamin D vs. placebo were also well balanced. The median duration of follow-up was 3.16 years (interquartile range, 2.83 to 3.38 years) and was not different between the two study arms. For the main trial, mean serum 25(OH)D_3_ concentrations at 3-year follow-up were higher among children randomised to receive vitamin D vs. placebo (104.3 vs. 64.7 nmol/L, respectively; mean difference 39.7 nmol/L, 95% CI for difference 37.6 to 41.9 nmol/L).

**Figure 1.**
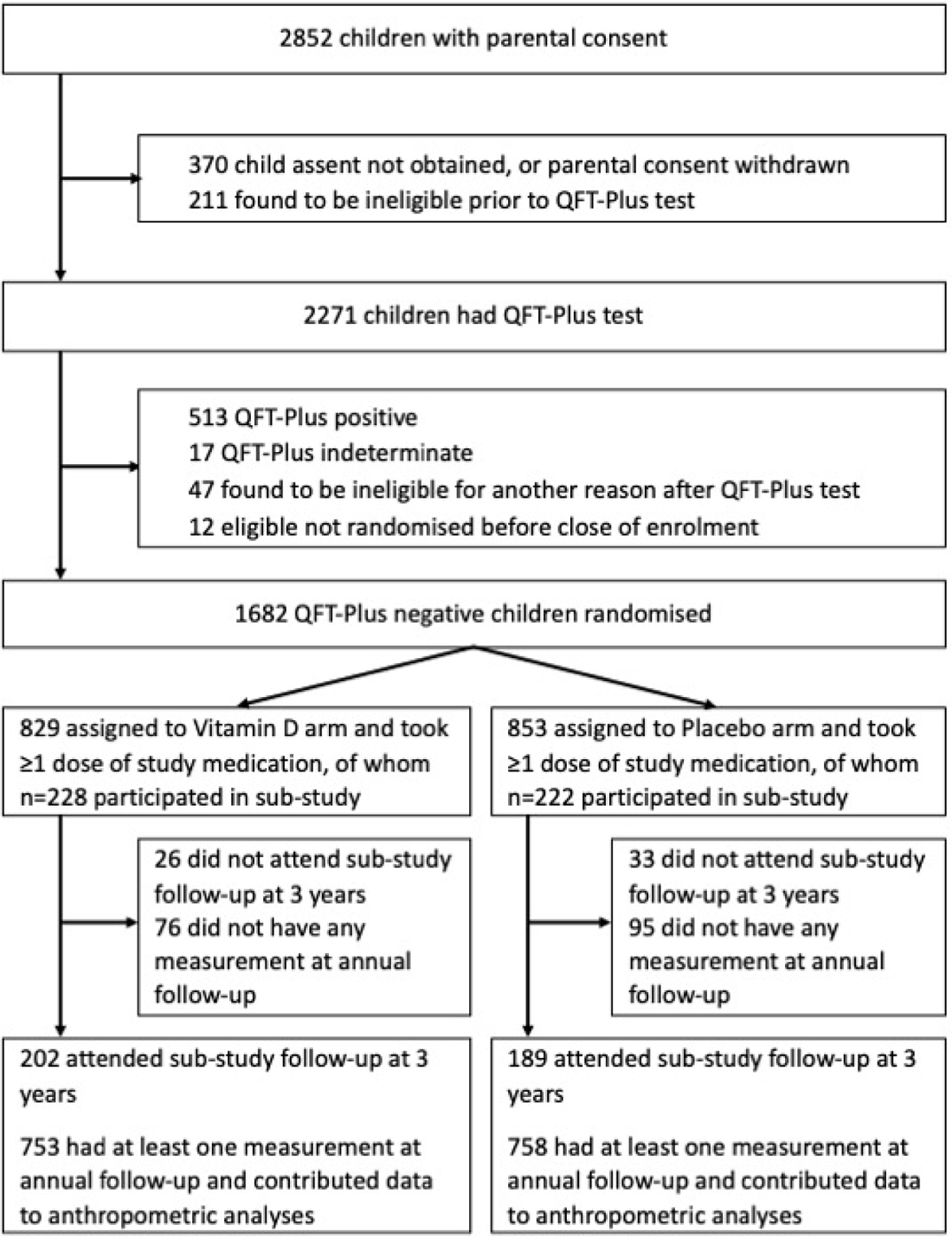
Participant Flow Diagram

**Table 1:**
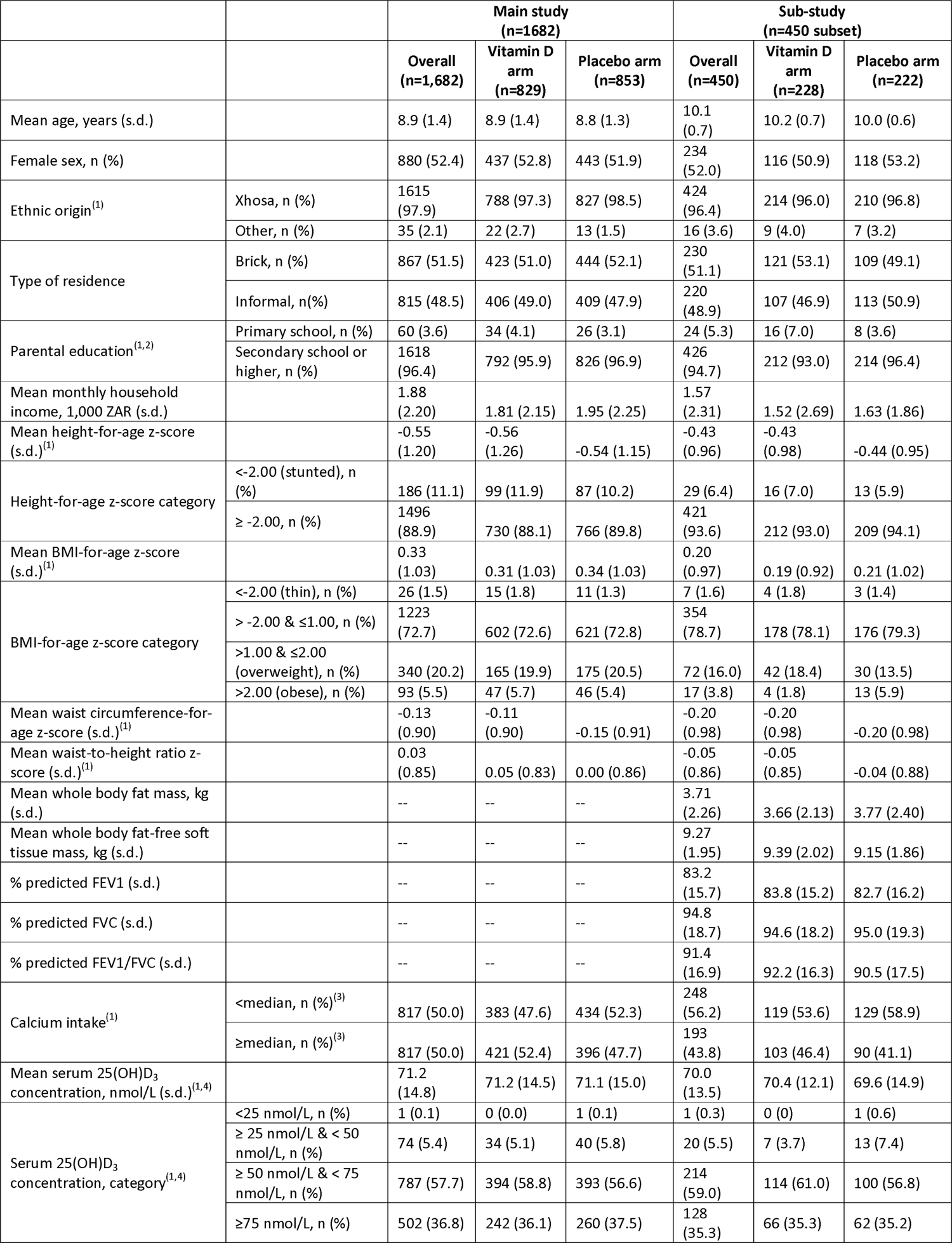
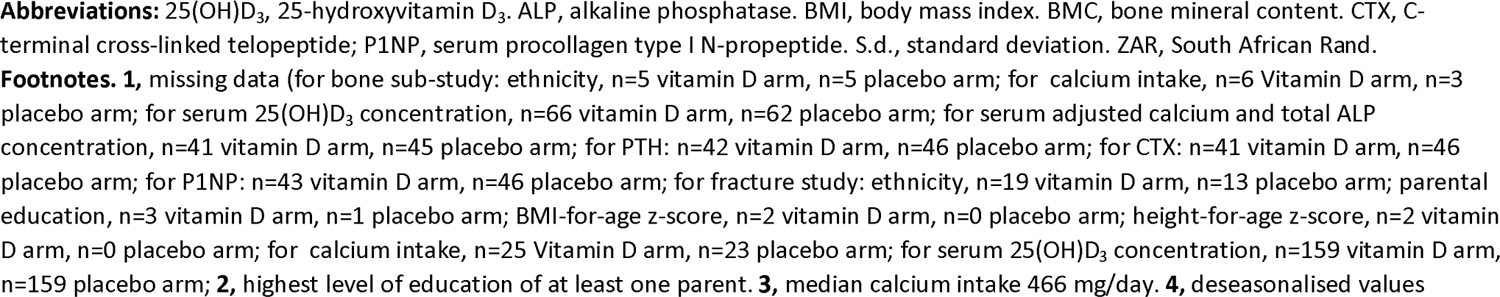
Participants’ baseline characteristics by allocation: main study and sub-study.

|  |  | Main study<br>(n=1682) |  |  | Sub-study<br>(n=450 subset) |  |  |
| --- | --- | --- | --- | --- | --- | --- | --- |
|  |  | Overall<br>(n=1,682) | Vitamin D<br>arm<br>(n=829) | Placebo arm<br>(n=853) | Overall<br>(n=450) | Vitamin D<br>arm<br>(n=228) | Placebo arm<br>(n=222) |
| Mean age, years (s.d.) |  | 8.9 (1.4) | 8.9 (1.4) | 8.8 (1.3) | 10.1<br>(0.7) | 10.2 (0.7) | 10.0 (0.6) |
| Female sex, n (%) |  | 880 (52.4) | 437 (52.8) | 443 (51.9) | 234<br>(52.0) | 116 (50.9) | 118 (53.2) |
| Ethnic origin<sup>(1)</sup> | Xhosa, n (%) | 1615<br>(97.9) | 788 (97.3) | 827 (98.5) | 424<br>(96.4) | 214 (96.0) | 210 (96.8) |
|  | Other, n (%) | 35 (2.1) | 22 (2.7) | 13 (1.5) | 16 (3.6) | 9 (4.0) | 7 (3.2) |
| Type of residence | Brick, n (%) | 867 (51.5) | 423 (51.0) | 444 (52.1) | 230<br>(51.1) | 121 (53.1) | 109 (49.1) |
|  | Informal, n(%) | 815 (48.5) | 406 (49.0) | 409 (47.9) | 220<br>(48.9) | 107 (46.9) | 113 (50.9) |
| Parental education<sup>(1,2)</sup> | Primary school, n (%) | 60 (3.6) | 34 (4.1) | 26 (3.1) | 24 (5.3) | 16 (7.0) | 8 (3.6) |
|  | Secondary school or<br>higher, n (%) | 1618<br>(96.4) | 792 (95.9) | 826 (96.9) | 426<br>(94.7) | 212 (93.0) | 214 (96.4) |
| Mean monthly household<br>income, 1,000 ZAR (s.d.) |  | 1.88<br>(2.20) | 1.81 (2.15) | 1.95 (2.25) | 1.57<br>(2.31) | 1.52 (2.69) | 1.63 (1.86) |
| Mean height-for-age z-score<br>(s.d.)<sup>(1)</sup> |  | -0.55<br>(1.20) | -0.56<br>(1.26) | -0.54 (1.15) | -0.43<br>(0.96) | -0.43<br>(0.98) | -0.44 (0.95) |
| Height-for-age z-score category | <-2.00 (stunted), n<br>(%) | 186 (11.1) | 99 (11.9) | 87 (10.2) | 29 (6.4) | 16 (7.0) | 13 (5.9) |
|  | ≥ -2.00, n (%) | 1496<br>(88.9) | 730 (88.1) | 766 (89.8) | 421<br>(93.6) | 212 (93.0) | 209 (94.1) |
| Mean BMI-for-age z-score<br>(s.d.)<sup>(1)</sup> |  | 0.33<br>(1.03) | 0.31 (1.03) | 0.34 (1.03) | 0.20<br>(0.97) | 0.19 (0.92) | 0.21 (1.02) |
| BMI-for-age z-score category | <-2.00 (thin), n (%) | 26 (1.5) | 15 (1.8) | 11 (1.3) | 7 (1.6) | 4 (1.8) | 3 (1.4) |
|  | > -2.00 & ≤1.00, n (%) | 1223<br>(72.7) | 602 (72.6) | 621 (72.8) | 354<br>(78.7) | 178 (78.1) | 176 (79.3) |
|  | >1.00 & ≤2.00<br>(overweight), n (%) | 340 (20.2) | 165 (19.9) | 175 (20.5) | 72 (16.0) | 42 (18.4) | 30 (13.5) |
|  | >2.00 (obese), n (%) | 93 (5.5) | 47 (5.7) | 46 (5.4) | 17 (3.8) | 4 (1.8) | 13 (5.9) |
| Mean waist circumference-for-age z-score (s.d.)<sup>(1)</sup> |  | -0.13<br>(0.90) | -0.11<br>(0.90) | -0.15 (0.91) | -0.20<br>(0.98) | -0.20<br>(0.98) | -0.20 (0.98) |
| Mean waist-to-height ratio z-score (s.d.)<sup>(1)</sup> |  | 0.03<br>(0.85) | 0.05 (0.83) | 0.00 (0.86) | -0.05<br>(0.86) | -0.05<br>(0.85) | -0.04 (0.88) |
| Mean whole body fat mass, kg<br>(s.d.) |  | -- | -- | -- | 3.71<br>(2.26) | 3.66 (2.13) | 3.77 (2.40) |
| Mean whole body fat-free soft<br>tissue mass, kg (s.d.) |  | -- | -- | -- | 9.27<br>(1.95) | 9.39 (2.02) | 9.15 (1.86) |
| % predicted FEV1 (s.d.) |  | -- | -- | -- | 83.2<br>(15.7) | 83.8 (15.2) | 82.7 (16.2) |
| % predicted FVC (s.d.) |  | -- | -- | -- | 94.8<br>(18.7) | 94.6 (18.2) | 95.0 (19.3) |
| % predicted FEV1/FVC (s.d.) |  | -- | -- | -- | 91.4<br>(16.9) | 92.2 (16.3) | 90.5 (17.5) |
| Calcium intake<sup>(1)</sup> | <median, n (%)<sup>(3)</sup> | 817 (50.0) | 383 (47.6) | 434 (52.3) | 248<br>(56.2) | 119 (53.6) | 129 (58.9) |
|  | ≥median, n (%)<sup>(3)</sup> | 817 (50.0) | 421 (52.4) | 396 (47.7) | 193<br>(43.8) | 103 (46.4) | 90 (41.1) |
| Mean serum 25(OH)D<sub>3</sub><br>concentration, nmol/L (s.d.)<sup>(1,4)</sup> |  | 71.2<br>(14.8) | 71.2 (14.5) | 71.1 (15.0) | 70.0<br>(13.5) | 70.4 (12.1) | 69.6 (14.9) |
| Serum 25(OH)D<sub>3</sub><br>concentration, category<sup>(1,4)</sup> | <25 nmol/L, n (%) | 1 (0.1) | 0 (0.0) | 1 (0.1) | 1 (0.3) | 0 (0) | 1 (0.6) |
|  | ≥ 25 nmol/L & < 50<br>nmol/L, n (%) | 74 (5.4) | 34 (5.1) | 40 (5.8) | 20 (5.5) | 7 (3.7) | 13 (7.4) |
|  | ≥ 50 nmol/L & < 75<br>nmol/L, n (%) | 787 (57.7) | 394 (58.8) | 393 (56.6) | 214<br>(59.0) | 114 (61.0) | 100 (56.8) |
|  | ≥75 nmol/L, n (%) | 502 (36.8) | 242 (36.1) | 260 (37.5) | 128<br>(35.3) | 66 (35.3) | 62 (35.2) |

**Abbreviations:** 25(OH)D<sub>3</sub>, 25-hydroxyvitamin D<sub>3</sub>. ALP, alkaline phosphatase. BMI, body mass index. BMC, bone mineral content. CTX, C-terminal cross-linked telopeptide; P1NP, serum procollagen type I N-propeptide. S.d., standard deviation. ZAR, South African Rand.
**Footnotes.** **1**, missing data (for bone sub-study: ethnicity, n=5 vitamin D arm, n=5 placebo arm; for calcium intake, n=6 Vitamin D arm, n=3 placebo arm; for serum 25(OH)D<sub>3</sub> concentration, n=66 vitamin D arm, n=62 placebo arm; for serum adjusted calcium and total ALP concentration, n=41 vitamin D arm, n=45 placebo arm; for PTH: n=42 vitamin D arm, n=46 placebo arm; for CTX: n=41 vitamin D arm, n=46 placebo arm; for P1NP: n=43 vitamin D arm, n=46 placebo arm; for fracture study: ethnicity, n=19 vitamin D arm, n=13 placebo arm; parental education, n=3 vitamin D arm, n=1 placebo arm; BMI-for-age z-score, n=2 vitamin D arm, n=0 placebo arm; height-for-age z-score, n=2 vitamin D arm, n=0 placebo arm; for calcium intake, n=25 Vitamin D arm, n=23 placebo arm; for serum 25(OH)D<sub>3</sub> concentration, n=159 vitamin D arm, n=159 placebo arm; **2**, highest level of education of at least one parent. **3**, median calcium intake 466 mg/day. **4**, deseasonalised values

### GROWTH OUTCOMES

Among participants in the main trial, allocation to vitamin D vs. placebo did not influence mean height-for-age z-scores at annual follow-up, either overall or within sub-groups defined by sex, calcium intake or baseline 25(OH)D concentration (Table 2: P values for interaction >0.05). Similarly, no effect of the intervention was seen on mean BMI-for-age z-scores (Table 3), waist circumference-for-age z-scores (Table S1) or waist-to-height ratio z-scores (Table S2), either overall or by sub-group (P values for interaction >0.05). Among sub-study participants, allocation to vitamin D vs. placebo did not influence fat mass or fat-free soft tissue mass (Table S3) or % predicted FEV1 or FEV1/FVC (Table S4), either overall or by sub-group. For the outcome of % predicted FVC, allocation to vitamin D vs. placebo did not have an effect overall, or within sub-groups defined by calcium intake or baseline 25(OH)D concentration; however, the P value for interaction associated with the sub-group analysis by sex (P=0.049) raised the possibility that this factor might modify the effect of vitamin D on % predicted FVC (Table S5).

**Table 2.** Mean height-for-age z-scores at annual follow-up by allocation, main study participants: overall and by sub-group.

|  |  | Follow-up time point | Vitamin D arm: mean value (s.d.) [n] | Placebo arm: mean value (s.d.) [n] | Adjusted mean difference (95% CI)<sup>1</sup> | P for timepoint | Overall P | P for interaction |
| --- | --- | --- | --- | --- | --- | --- | --- | --- |
| Overall |  | 1 year | -0.22 (1.12) [664] | -0.15 (1.07) [661] | -0.06 (-0.17 to 0.06) | 0.33 | 0.22 | -- |
|  |  | 2 years | -0.14 (1.05) [613] | -0.06 (1.01) [606] | -0.04 (-0.16 to 0.07) | 0.46 |  |  |
|  |  | 3 years | -0.26 (1.04) [670] | -0.14 (1.05) [691] | -0.08 (-0.19 to 0.03) | 0.17 |  |  |
| By sex | Boys | 1 year | -0.31 (1.14) [304] | -0.27 (1.06) [306] | -0.02 (-0.17 to 0.14) | 0.85 | 0.42 | 0.94 |
|  |  | 2 years | -0.21 (1.11) [285] | -0.19 (0.95) [283] | 0.04 (-0.13 to 0.20) | 0.67 |  |  |
|  |  | 3 years | -0.34 (1.07) [310] | -0.28 (1.00) [326] | -0.03 (-0.19 to 0.13) | 0.69 |  |  |
|  | Girls | 1 year | -0.15 (1.09) [360] | -0.04 (1.07) [355] | -0.10 (-0.25 to 0.06) | 0.21 | 0.34 |  |
|  |  | 2 years | -0.08 (0.99) [328] | 0.05 (1.06) [323] | -0.12 (-0.28 to 0.04) | 0.14 |  |  |
|  |  | 3 years | -0.19 (1.02) [360] | -0.02 (1.08) [365] | -0.13 (-0.28 to 0.03) | 0.11 |  |  |
| By calcium intake<sup>2</sup> | <median | 1 year | -0.16 (1.05) [314] | -0.10 (1.02) [331] | -0.04 (-0.18 to 0.11) | 0.64 | 0.02 | 0.07 |
|  |  | 2 years | -0.17 (0.96) [288] | -0.05 (0.96) [310] | -0.04 (-0.19 to 0.11) | 0.64 |  |  |
|  |  | 3 years | -0.28 (0.99) [312] | -0.15 (0.99) [362] | -0.08 (-0.23 to 0.07) | 0.29 |  |  |
|  | ≥median | 1 year | -0.25 (1.16) [327] | -0.18 (1.13) [314] | -0.05 (-0.21 to 0.12) | 0.60 | 0.72 |  |
|  |  | 2 years | -0.07 (1.12) [307] | -0.06 (1.06) [279] | -0.03 (-0.20 to 0.14) | 0.76 |  |  |
|  |  | 3 years | -0.20 (1.09) [337] | -0.13 (1.11) [311] | -0.06 (-0.22 to 0.11) | 0.50 |  |  |
| By baseline 25(OH)D concentration<sup>3</sup> | <75 nmol/L | 1 year | -0.23 (1.13) [349] | -0.03 (1.11) [332] | -0.17 (-0.33 to -0.01) | 0.04 | 0.53 | 0.50 |
|  |  | 2 years | -0.14 (1.11) [325] | 0.05 (1.06) [323] | -0.12 (-0.28 to 0.04) | 0.15 |  |  |
|  |  | 3 years | -0.25 (1.10) [340] | -0.06 (1.07) [359] | -0.15 (-0.31 to 0.01) | 0.07 |  |  |
|  | ≥75 nmol/L | 1 year | -0.22 (1.15) [187] | -0.25 (0.98) [201] | 0.02 (-0.17 to 0.22) | 0.82 | 0.15 |  |
|  |  | 2 years | -0.17 (1.00) [171] | -0.08 (0.96) [179] | -0.07 (-0.26 to 0.13) | 0.50 |  |  |
|  |  | 3 years | -0.30 (1.00) [196] | -0.18 (0.99) [206] | -0.06 (-0.25 to 0.13) | 0.53 |  |  |

**Abbreviations:** 25(OH)D, 25-hydroxyvitamin D. CI, confidence interval. S.d., standard deviation. n, number.
**Footnotes.** 1, adjusted for baseline value and school of attendance. 2, median calcium intake 466 mg/day. 3, deseasonalised values.

**Table 3.** Mean BMI-for-age z-scores at annual follow-up by allocation, main study participants: overall and by sub-group.

|  |  |  | Vitamin D arm:<br>mean value (s.d.)<br>[n] | Placebo arm:<br>mean value (s.d.)<br>[n] | Adjusted mean<br>difference (95% CI)<sup>1</sup> | P for<br>timepoint | Overall<br>P | P for<br>interaction |
| --- | --- | --- | --- | --- | --- | --- | --- | --- |
| Overall |  | 1 year | 0.16 (1.03) [664] | 0.17 (1.02) [661] | -0.02 (-0.14 to 0.09) | 0.68 | 0.86 | -- |
|  |  | 2 years | 0.15 (1.41) [613] | 0.25 (1.21) [606] | -0.09 (-0.20 to 0.03) | 0.14 |  |  |
|  |  | 3 years | 0.17 (1.07) [669] | 0.21 (1.17) [691] | -0.04 (-0.16 to 0.07) | 0.45 |  |  |
| By sex | Boys | 1 year | 0.08 (0.99) [304] | -0.00 (0.99) [306] | 0.09 (-0.08 to 0.27) | 0.30 | 0.59 | 0.29 |
|  |  | 2 years | 0.00 (1.68) [285] | -0.04 (1.30) [283] | 0.04 (-0.14 to 0.22) | 0.64 |  |  |
|  |  | 3 years | -0.02 (1.04) [309] | -0.12 (1.20) [326] | 0.09 (-0.09 to 0.26) | 0.33 |  |  |
|  | Girls | 1 year | 0.23 (1.06) [360] | 0.32 (1.02) [355] | -0.14 (-0.28 to 0.00) | 0.06 | 0.26 |  |
|  |  | 2 years | 0.28 (1.10) [328] | 0.50 (1.06) [323] | -0.21 (-0.36 to -0.06) | 0.00 |  |  |
|  |  | 3 years | 0.33 (1.06) [360] | 0.50 (1.07) [365] | -0.17 (-0.32 to -0.03) | 0.02 |  |  |
| By calcium<br>intake<sup>2</sup> | <median | 1 year | 0.15 (1.01) [314] | 0.15 (1.03) [331] | -0.01 (-0.17 to 0.14) | 0.86 | 0.16 | 0.08 |
|  |  | 2 years | 0.18 (1.08) [288] | 0.21 (1.34) [310] | -0.03 (-0.19 to 0.13) | 0.75 |  |  |
|  |  | 3 years | 0.21 (1.07) [312] | 0.20 (1.14) [362] | 0.01 (-0.14 to 0.17) | 0.87 |  |  |
|  | ≥median | 1 year | 0.18 (1.04) [327] | 0.22 (1.01) [314] | -0.04 (-0.21 to 0.13) | 0.61 | 0.27 |  |
|  |  | 2 years | 0.13 (1.67) [307] | 0.31 (1.07) [279] | -0.15 (-0.32 to 0.03) | 0.10 |  |  |
|  |  | 3 years | 0.15 (1.07) [336] | 0.22 (1.23) [311] | -0.09 (-0.26 to 0.08) | 0.31 |  |  |
| By baseline<br>25(OH)D<br>concentration<sup>3</sup> | <75<br>nmol/L | 1 year | 0.30 (1.03) [349] | 0.28 (0.98) [332] | 0.03 (-0.14 to 0.19) | 0.76 | 0.31 | 0.31 |
|  |  | 2 years | 0.22 (1.64) [325] | 0.31 (1.33) [323] | -0.07 (-0.24 to 0.10) | 0.41 |  |  |
|  |  | 3 years | 0.25 (1.07) [340] | 0.34 (1.08) [359] | -0.09 (-0.25 to 0.08) | 0.31 |  |  |
|  | ≥75<br>nmol/L | 1 year | 0.02 (0.97) [187] | 0.09 (1.02) [201] | -0.06 (-0.25 to 0.13) | 0.57 | 0.58 |  |
|  |  | 2 years | 0.01 (1.05) [171] | 0.18 (1.00) [179] | -0.11 (-0.30 to 0.09) | 0.28 |  |  |
|  |  | 3 years | 0.09 (1.00) [196] | 0.06 (1.17) [206] | 0.02 (-0.17 to 0.21) | 0.87 |  |  |

**Abbreviations:** 25(OH)D, 25-hydroxyvitamin D. CI, confidence interval. S.d., standard deviation. n, number.
**Footnotes.** 1, adjusted for baseline value and school of attendance. 2, median calcium intake 466 mg/day. 3, deseasonalised values.

### DEVELOPMENTAL OUTCOMES

Among sub-study participants, no inter-arm differences were seen in mean Tanner scores for pubic hair (males and females), external genitalia (males only), proportion menstruating (females only), mean age at menarche (females only), or breast development (females only), either overall or within sub-groups defined by calcium intake or baseline 25(OH)D concentration (Table 4).

**Table 4.** End-study pubertal development indices by allocation, sub-study participants: overall and by sub-group.

|  |  |  |  | Vitamin D arm:<br>mean value<br>(s.d.) [n] | Placebo arm:<br>mean value (s.d.)<br>[n] | Adjusted mean<br>difference (95% CI)<sup>1</sup> | P | P for<br>interaction |
| --- | --- | --- | --- | --- | --- | --- | --- | --- |
| Males | Pubic hair,<br>mean Tanner<br>score (s.d.)<br>[n] | Overall |  | 3.15 (0.91) [96] | 2.81 (0.99) [90] | 0.25 (-0.02 to 0.51) | 0.07 | -- |
|  |  | Calcium<br>intake<sup>2</sup> | <median | 3.13 (0.93) [46] | 2.73 (1.14) [52] | 0.29 (-0.10 to 0.67) | 0.14 | 0.39 |
|  |  |  | ≥median | 3.15 (0.90) [48] | 2.92 (0.75) [38] | 0.16 (-0.19 to 0.52) | 0.37 |  |
|  |  | Baseline<br>25(OH)D<br>concentration<sup>3</sup> | <75<br>nmol/L | 3.11 (0.92) [44] | 2.88 (0.92) [42] | 0.07 (-0.28 to 0.41) | 0.70 | 0.39 |
|  |  |  | ≥75<br>nmol/L | 3.18 (0.94) [34] | 2.70 (1.18) [30] | 0.43 (-0.07 to 0.93) | 0.09 |  |
|  | External<br>genitalia,<br>mean Tanner<br>score (s.d.)<br>[n] | Overall |  | 3.19 (0.91) [96] | 2.90 (0.99) [90] | 0.17 (-0.09 to 0.43) | 0.19 | -- |
|  |  | Calcium<br>intake<sup>2</sup> | <median | 3.28 (0.89) [46] | 2.81 (1.10) [52] | 0.35 (-0.01 to 0.71) | 0.06 | 0.07 |
|  |  |  | ≥median | 3.10 (0.93) [48] | 3.03 (0.82) [38] | -0.04 (-0.40 to 0.33) | 0.84 |  |
|  |  | Baseline<br>25(OH)D<br>concentration<sup>3</sup> | <75<br>nmol/L | 3.27 (0.82) [44] | 2.90 (0.91) [42] | 0.22 (-0.10 to 0.54) | 0.18 | 0.81 |
|  |  |  | ≥75<br>nmol/L | 3.15 (1.02) [34] | 2.87 (1.17) [30] | 0.30 (-0.17 to 0.77) | 0.21 |  |
| Females | Pubic hair,<br>mean Tanner<br>score (s.d.)<br>[n] | Overall |  | 3.10 (0.98) [105] | 3.11 (0.98) [99] | -0.09 (-0.35 to 0.17) | 0.50 | -- |
|  |  | Calcium<br>intake<sup>2</sup> | <median | 3.15 (0.98) [47] | 3.15 (1.01) [46] | 0.01 (-0.38 to 0.40) | 0.96 | 0.60 |
|  |  |  | ≥median | 3.11 (0.92) [54] | 3.10 (0.97) [50] | -0.23 (-0.56 to 0.10) | 0.17 |  |
|  |  | Baseline<br>25(OH)D<br>concentration<sup>3</sup> | <75<br>nmol/L | 3.14 (0.90) [59] | 3.13 (0.88) [55] | -0.03 (-0.35 to 0.28) | 0.83 | 0.94 |
|  |  |  | ≥75<br>nmol/L | 2.96 (1.22) [26] | 3.04 (1.08) [24] | -0.14 (-0.74 to 0.46) | 0.65 |  |
|  | Breast<br>development,<br>mean Tanner<br>score (s.d.)<br>[n] | Overall |  | 3.15 (0.87) [105] | 3.10 (0.91) [99] | -0.03 (-0.26 to 0.20) | 0.80 | -- |
|  |  | Calcium<br>intake<sup>2</sup> | <median | 3.15 (0.91) [47] | 3.22 (0.92) [46] | -0.06 (-0.41 to 0.29) | 0.74 | 0.93 |
|  |  |  | ≥median | 3.20 (0.81) [54] | 3.04 (0.90) [50] | -0.05 (-0.36 to 0.26) | 0.76 |  |
|  |  | Baseline<br>25(OH)D<br>concentration<sup>3</sup> | <75<br>nmol/L | 3.27 (0.85) [59] | 3.07 (0.86) [55] | 0.15 (-0.16 to 0.45) | 0.34 | 0.48 |
|  |  |  | ≥75<br>nmol/L | 3.08 (0.93) [26] | 3.08 (0.97) [24] | -0.08 (-0.55 to 0.38) | 0.73 |  |
|  | Proportion<br>menstruating | Overall |  | 67/105 (63.81) | 60/99 (60.61) | 0.95 (0.51 to 1.77) | 0.86 | -- |
|  |  | Calcium<br>intake<sup>2</sup> | <median | 28/47 (59.57) | 26/46 (56.52) | 1.33 (0.50 to 3.56) | 0.57 | 0.24 |
|  |  |  | ≥median | 38/54 (70.37) | 34/50 (68.00) | 0.56 (0.21 to 1.52) | 0.26 |  |
|  |  | Baseline<br>25(OH)D<br>concentration<sup>3</sup> | <75<br>nmol/L | 43/59 (72.88) | 35/55 (63.64) | 1.39 (0.59 to 3.28) | 0.45 | 0.59 |
|  |  |  | ≥75<br>nmol/L | 16/26 (61.54) | 15/24 (62.50) | 0.90 (0.26 to 3.15) | 0.87 |  |
|  | Mean age at<br>menarche,<br>years (s.d.)<br>[n] | Overall |  | 11.94 (0.93) [66] | 11.86 (0.66) [58] | -0.03 (-0.31 to 0.25) | 0.84 | -- |
|  |  | Calcium<br>intake<sup>2</sup> | <median | 11.89 (0.83) [28] | 11.71 (0.69) [24] | 0.12 (-0.26 to 0.49) | 0.54 | 0.32 |
|  |  |  | ≥median | 11.97 (1.01) [37] | 11.97 (0.63) [34] | -0.11 (-0.51 to 0.29) | 0.58 |  |
|  |  | Baseline<br>25(OH)D<br>concentration<sup>3</sup> | <75<br>nmol/L | 12.00 (0.88) [42] | 11.88 (0.69) [34] | 0.04 (-0.31 to 0.39) | 0.82 | 0.64 |
|  |  |  | ≥75<br>nmol/L | 11.88 (1.15) [16] | 11.93 (0.62) [14] | -0.14 (-0.73 to 0.44) | 0.63 |  |

**Abbreviations:** 25(OH)D, 25-hydroxyvitamin D. CI, confidence interval. S.d., standard deviation. n, number.
**Footnotes.** 1, adjusted for school of attendance. 2, median calcium intake 466 mg/day. 3, deseasonalised values.

### ADVERSE EVENTS

Incidence of adverse events by trial arm has been reported elsewhere.^18^ No serious events arising in the trial were adjudged related to administration of vitamin D or placebo.

## DISCUSSION

We report findings from the first randomised controlled trial to investigate effects of vitamin D supplementation on growth and developmental indices in schoolchildren of Black African ancestry. Administration of oral vitamin D supplementation at a dose of 10,000 IU per week for 3 years was effective in boosting vitamin D status, but it did not impact linear growth, body mass index, body composition, spirometric lung volumes or self-assessed pubertal development, either overall or in sub-groups defined by sex, calcium intake or baseline vitamin D status.

Null findings from the current study contrast with those from observational studies reporting independent associations between vitamin D deficiency and reduced lean mass,^8^ slower linear growth,^7^ childhood obesity^9–11^ and precocious puberty.^12^ However, they are consistent with those of the only other phase 3 RCT to investigate effects of vitamin D on growth and development in school-age children that we are aware of, which reported no effect of weekly vitamin D supplementation on growth or development among Mongolian schoolchildren aged 6-13 years at baseline.^17^ Contrasting findings from observational studies vs. clinical trials may reflect the fact that the former are more susceptible to effects of confounding or reverse causality. Although we observed limited evidence (P for interaction 0.049) to support the hypothesis that effects of vitamin D on FVC might be modified by sex, this finding may have arisen because of type 1 error, given the multiplicity of analyses conducted.

Our study has several strengths. The intervention was sustained (3 years), allowing ample time for any effects of vitamin D supplementation on outcomes of interest to manifest. Moreover, the intervention was effective in elevating serum 25(OH)D concentrations, reflecting adequacy of the dosing regimen employed as well as good adherence resulting from directly observed administration of weekly supplements in schools during termtime. Our large sample size and low rates of loss to follow-up maximised power to detect modest effects of the intervention, particularly for outcomes that were assessed in the main trial population. Furthermore, we assessed a broad range of anthropometric and developmental outcomes, that included use of DXA, the gold standard investigation for assessment of body composition.

Our study also has some limitations. Baseline prevalence of vitamin D deficiency was low, perhaps reflecting plentiful exposure to sunshine in the study setting.^22^ Our results cannot therefore be generalised to settings where vitamin D deficiency is common. However, we highlight that results from our trial in Mongolia (where baseline prevalence of vitamin D deficiency was much higher than we observed in the current study) were also null for outcomes of linear growth and body mass index.^17^ Accordingly, null results from the current trial cannot be explained simply by the relatively high baseline vitamin D status of participants in Cape Town. A second potential limitation relates to the fact that pubertal status was assessed by participants themselves, rather than by health professionals, whose judgement may be more objective. However, the double-blind placebo-controlled trial design of our study will have distributed any resultant imprecision equally between study arms, ensuring that bias was not introduced. Moreover, pubertal self-assessment has previously been done by adolescent participants in another South African cohort study, and shown to be reliable.^25^

In conclusion, we report that oral vitamin D supplementation at a dose of 10,000 IU/week for 3 years was effective in elevating serum 25(OH)D concentrations in schoolchildren of Black African ancestry living in Cape Town, South Africa. However, this was not associated with any effects on linear growth, body habitus, spirometric outcomes or pubertal development.

## Data Availability

Anonymised data may be requested from the corresponding author to be shared subject to terms of research ethics committee approval.

## ACKNOWLEDGEMENTS

This research was funded by the UK Medical Research Council (refs MR/R023050/1 and MR/M026639/1, both awarded to ARM). RJW was supported by Wellcome (104803, 203135). He also received support from the Francis Crick Institute which is funded by Cancer Research UK (FC2112), the UK Medical Research Council (FC2112) and Wellcome (FC2112). We thank all the children who participated in the trial, and their parents / guardians; members of the Independent Data Monitoring Committee (Prof Guy Thwaites, Oxford University Clinical Research Unit, Ho Chi Minh City, Vietnam [Chair]; Prof John Pettifor, University of the Witwatersrand, Johannesburg, South Africa; and Prof Sarah Walker, MRC Clinical Trials Unit, London, UK); and members of the Trial Steering Committee (Prof Beate Kampmann, London School of Hygiene and Tropical Medicine, London, UK [Chair]; Prof Ashraf Coovadia, University of the Witwatersrand, Johannesburg, South Africa; Dr Karen Jennings, City Health, Cape Town, South Africa; Dr Robin Dyers, Department of Health, Western Cape Government, Cape Town, South Africa; and Dr Guy de Bruyn, Sanofi Pasteur, Swiftwater PA USA). For the purposes of open access the author has applied a CC-BY public copyright to any author-accepted manuscript arising from this submission.

## CONTRIBUTORS

ARM conceived the study. KM, LKM, AKC, JN, CC, NCH, RLH, RJW, LGB and ARM contributed to study design and protocol development. KM led on trial implementation, with support from JS, CD, DAJ, JN, LGB and ARM. LKM and AEM oversaw performance of DXA scans. JCYT and WDF performed and supervised the conduct of biochemical assays. NW, RLH and ARM drafted the statistical analysis plan. DAJ, KM, JS, NW, WM and CD managed data. NW accessed, verified and analysed the data underlying the study. ARM and DAJ wrote the first draft of the trial report. All authors made substantive comments thereon and approved the final version for submission.

