## Supplementary Appendix for "Influence of vitamin D supplementation on growth, body composition, pubertal development and spirometry in South African schoolchildren: a randomised controlled trial (ViDiKids)"

**Supplemental Material**

### **Supplementary Methods**

#### ***Enrolment***

Parents or legal guardians were invited to provide written informed consent for their child to participate in the main trial during a home visit, unless their child was eligible for the sub-study, in which case they were invited to provide written informed consent for their child to participate in both the main trial and the sub-study until a total of 450 sub-study participants were randomised. If parents / legal guardians consented, they were asked to provide details of their child’s dietary intake of foods containing vitamin D and calcium in the previous month, which were captured on an electronic case report form as previously reported.^19^ Their children were then invited to provide written assent to participate in the main trial +/- the sub-study (if eligible) at a school-based visit. If they agreed, a clinically trained member of the study team screened them for symptoms and signs of rickets. For all participants, a blood sample was taken for a QFT-Plus assay and separation and storage of serum for determination of 25(OH)D concentrations as described below. Participants were reviewed when baseline QFT-Plus results were available. Those with a positive QFT-Plus result were excluded from the trial and screened for active TB. Those with an indeterminate QFT-Plus result were excluded from the trial without screening for active TB. Those with a negative QFT-Plus result were deemed eligible to participate and underwent measurement of weight (using a digital floor scale, Charder Medical, Taichung City, Taiwan), height (using a portable HM200P stadiometer, Charder Medical) and waist circumference (using a measuring tape). Sub-study participants also underwent spirometry according to ERS/ATS standards^20^ using a portable spirometer (Carefusion, San Diego, CA) and measurement of body composition by dual energy x-ray absorptiometry (DXA) as described below.

#### ***Randomisation and blinding***

Eligible participants were individually randomised to receive a weekly capsule containing vitamin D_3_ or placebo for three years, with a one-to-one allocation ratio. Randomisation was stratified by school of attendance (a potential predictor of risk of QFT conversion) as follows. Prior to the start of recruitment, Mrs Claire Chan (Independent statistician, Pragmatic Clinical Trials Unit, Queen Mary University of London) prepared two school randomisation lists, each comprising 20 pairs of 2-letter randomisation codes, with each pair allocated to a single school identifier (e.g. School 1 was allocated codes ‘AC’ and ‘MO’, School 2 was allocated codes ‘BD’ and ‘NP’). One 2-letter randomisation code within each pair was then randomly assigned to the vitamin D arm of the trial, and the other was assigned to the placebo arm, using a computer-generated random sequence (e.g. ‘AC’ and ‘BD’ were assigned to ‘vitamin D’, ‘MO’ and ‘NP’ were assigned to placebo). Mrs Chan also prepared 40 separate participant randomisation lists (one for each potential participating school). Each of these participant randomisation lists comprised 999 5-digit numbers, each consisting of a 2-digit school identifier from 01 to 25 that was constant for, and unique to, each list, followed by a 3-digit participant identifier from 001 to 999 (e.g. 01-001, 01-002, 02-001, 02-002). These 5-digit numbers were then randomly assigned to one or other of the 2-letter randomisation codes allocated to that school in blocks of ten, using a computer-generated random sequence (e.g. for School 1, sequence was 01-001-MO, 01-002-AC, etc.; for School 2, sequence was 02-001-BD, 02-002-BD, etc.).

Active and placebo capsules were shipped from Tishcon Corporation (Westbury, NY, USA) to Lekoko Pharmaceutical Management Consultancy (Johannesburg, South Africa) in boxes that were labelled ‘vitamin D’ or ‘placebo’ according to their contents. On arrival, these capsules were packed into bottles, each containing either 150 vitamin D capsules or 150 placebo capsules. The school randomisation list was then used to label these bottles with 2-letter randomisation codes according to their contents (i.e. bottles containing vitamin D capsules were labelled with ‘AC’, ‘BD’ or another 2-letter code assigned to the vitamin D arm of the trial, while bottles containing placebo capsules were labelled ‘MO’, ‘NP’ or another 2-letter code assigned to the placebo arm of the trial) until sufficient capsules for 23 schools had been bottled and labelled. Bottling and labelling was performed under the supervision of Mr Bobby Hamman (Lekoko PMC), with independent monitoring performed by staff from OnQ Contract Research Organisation; none of these individuals was involved with data collection. Children screened at each school were assigned consecutive 5-digit numbers at enrolment by study field workers, and if they were subsequently found to be eligible for randomisation (i.e. if their baseline QFT-Plus result was negative) then the participant randomisation list for their school of attendance was used to determine their allocation, i.e. they received study medication from bottles labelled with the 2-letter code linked to their 5-digit ID in the participant randomization list for the duration of the trial. For example, participant 01-001 would receive study medication labelled ‘MO’ (i.e. placebo), 01-002 would receive medication labelled ‘AC’ (i.e. vitamin D), 02-001 would receive medication labelled ‘BD’ (vitamin D), and 02-002 would receive medication labelled ‘BD’ (vitamin D) throughout the trial. Copies of the school randomisation list were held by Mrs Chan and by members of the DSMB. Neither participants nor trial staff had access to it, and treatment allocation was concealed from participants, care providers and all trial staff (including senior investigators and those assessing outcomes) so that the double-blind was maintained. The school randomisation lists were made available to Dr Neil Walker (trial statistician) following completion of the trial, who used them to un-blind allocation and to analyse the trial: he was not therefore masked to group assignment during statistical analysis.

#### ***Intervention***

Study medication comprised a 3-year course of weekly soft gel capsules manufactured by the Tishcon Corporation (Westbury, NY, USA), containing either 0.25 mg (10,000 international units) cholecalciferol (vitamin D_3_) in olive oil (intervention arm) or olive oil without any vitamin D_3_ content (placebo arm). Active and placebo capsules had identical appearance and taste. Capsules were taken under direct observation of study staff during school term time. Further details of administration of study medication are provided in Supplementary Material. During summer holidays (8 weeks), packs containing 8 doses of study medication were provided for administration by parents, together with a participant diary. Following shorter school holidays (≤4 weeks), and/or if participants missed one or more doses of study medication during term time, up to 4 ‘catch-up’ doses were administered at the first weekly visit attended following the missed dose(s). During the initial national lockdown for COVID-19 in South Africa (27^th^ March to 1^st^ May 2020), participants did not receive any study medication. During subsequent school closures due to COVID-19, two rounds of 8-week holiday packs were provided to participants, which were sufficient to cover their requirements until schools re-opened.

#### ***Laboratory Assessments***

Biochemical analyses were performed at the Bioanalytical Facility, University of East Anglia (Norwich, UK) according to manufacturers’ instructions and under Good Clinical and Laboratory Practice conditions. Serum concentrations of 25(OH)D_3_ were measured using liquid chromatography tandem mass spectrometry (LC-MS/MS) as previously described.^22^ 25(OH)D_3_ was calibrated using standard reference material SRM972a from the National Institute of Science and Technology (NIST), and the assay showed linearity between 0 and 200 nmol/L. The inter/intra-assay coefficient of variation (CV) across the assay range was ≤9%, and the lower limit of quantification was 0.1 nmol/L. The assay showed <6% accuracy bias against NIST reference method on the vitamin D external quality assessment (DEQAS) scheme (<http://www.deqas.org/>; accessed on 30^th^ November 2022). QFT-Plus assays were performed by the Bio Analytical Research Corporation South Africa (Johannesburg, South Africa) according to the manufacturer’s instructions.

#### ***Statistical Analyses***

Statistical analyses were performed using Stata software (Version 17.0; StataCorp, College Station, Texas, United States) according to intention to treat. Anthropometric measurements were used to compute age- and sex-adjusted z-scores for height, BMI, waist circumference and waist-to-height ratio, using the WHO 2007 Lambda, Mu and Sigma method.^23^ Effects of treatment on the above Z-transformed outcomes were estimated by fitting allocation to vitamin D vs. placebo as the sole fixed effect in a mixed effects linear regression model with a random effect for repeated assessments of each individual participant (at baseline and years 1, 2 and 3 of follow-up) and a random effect of school of attendance, with results reported as treatment differences with 95% confidence intervals. Effects of treatment on DXA and spirometric outcomes measured at 3-year follow-up were analysed in each case using a multi-level mixed model with an adjustment for analogous baseline score and a random effect for school. Tanner metrics on pubertal development were analysed in a similar fashion, but restricted to the applicable sex and without baseline adjustment. Binary outcomes measured at 3-year follow-up only were analysed using a mixed-effects logistic regression model, with treatment allocation as the sole fixed effect and school attended as a random intercept. Pre-specified sub-group analyses were conducted to determine whether the effect of vitamin D supplementation was modified by sex (male vs. female), baseline deseasonalised 25(OH)D_3_ concentration (<75 vs. ≥75 nmol/L, calculated using a sinusoidal model as previously described)^24^ and calcium intake (< vs. ≥ median value of 466 mg/day, calculated from a dietary intake questionnaire as previously described).^19^ These were performed by repeating efficacy analyses with the inclusion of an interaction term between allocation (to vitamin D vs. placebo) and each posited effect-modifier with presentation of the P-value associated with this interaction term. Effect of sex as a subgroup was not considered in relation to analyses of Tanner outcomes, as these are sex-specific. Given the number of potential effect modifiers and secondary outcome measures these analyses are considered exploratory. Interim safety assessments, where Independent Data Monitoring Committee (IDMC) members reviewed accumulating serious adverse event data, were performed at 6-monthly intervals. At each review, the IDMC recommended continuation of the trial. No interim efficacy analysis was performed.

### **Table S1.** Mean waist circumference-for-age z-scores at annual follow-up by allocation, main trial participants: overall and by sub-group

|  |  |  | **Vitamin D: mean value (s.d.) [n]** | **Placebo: mean value (s.d.) [n]** | **Adjusted mean difference (95% CI)^1^** | **P for timepoint** | **P for trend** | **P for interaction**  **(treatment*subgroup)** |
| --- | --- | --- | --- | --- | --- | --- | --- | --- |
| Overall |  | 1 year | -0.03 (0.89) [664] | -0.00 (0.87) [660] | -0.03 (-0.12 to 0.06) | 0.47 | 0.01 |  |
|  |  | 2 years | 0.19 (0.87) [612] | 0.24 (0.87) [608] | -0.06 (-0.15 to 0.04) | 0.24 |  |  |
|  |  | 3 years | 0.15 (0.83) [667] | 0.21 (0.88) [686] | -0.07 (-0.16 to 0.02) | 0.12 |  |  |
| By sex | Boys | 1 year | -0.14 (0.84) [304] | -0.20 (0.88) [305] | 0.06 (-0.07 to 0.19) | 0.39 | 0.07 | 0.93 |
|  |  | 2 years | 0.11 (0.84) [286] | 0.06 (0.88) [284] | 0.05 (-0.08 to 0.18) | 0.47 |  |  |
|  |  | 3 years | 0.03 (0.83) [309] | 0.03 (0.85) [325] | -0.02 (-0.15 to 0.11) | 0.78 |  |  |
|  | Girls | 1 year | 0.06 (0.92) [360] | 0.17 (0.82) [355] | -0.12 (-0.24 to 0.00) | 0.05 | 0.04 |  |
|  |  | 2 years | 0.26 (0.88) [326] | 0.41 (0.84) [324] | -0.15 (-0.28 to -0.03) | 0.01 |  |  |
|  |  | 3 years | 0.25 (0.82) [358] | 0.37 (0.86) [361] | -0.13 (-0.25 to -0.01) | 0.04 |  |  |
| By calcium intake^2^ | <median | 1 year | -0.04 (0.89) [314] | -0.05 (0.86) [331] | 0.00 (-0.12 to 0.13) | 0.96 | 0.02 | 0.64 |
|  |  | 2 years | 0.19 (0.86) [289] | 0.19 (0.89) [312] | 0.00 (-0.12 to 0.13) | 0.94 |  |  |
|  |  | 3 years | 0.16 (0.81) [310] | 0.21 (0.85) [358] | -0.03 (-0.16 to 0.09) | 0.62 |  |  |
|  | ≥median | 1 year | -0.01 (0.89) [327] | 0.06 (0.89) [313] | -0.07 (-0.20 to 0.06) | 0.28 | 0.10 |  |
|  |  | 2 years | 0.20 (0.87) [305] | 0.30 (0.86) [279] | -0.10 (-0.23 to 0.03) | 0.14 |  |  |
|  |  | 3 years | 0.14 (0.87) [337] | 0.22 (0.91) [310] | -0.11 (-0.24 to 0.02) | 0.10 |  |  |
| By baseline 25(OH)D concentration^3^ | <75 nmol/L | 1 year | 0.09 (0.90) [349] | 0.08 (0.87) [332] | 0.02 (-0.10 to 0.15) | 0.72 | 0.08 | 0.68 |
|  |  | 2 years | 0.26 (0.89) [324] | 0.31 (0.88) [324] | -0.02 (-0.15 to 0.11) | 0.75 |  |  |
|  |  | 3 years | 0.21 (0.85) [338] | 0.29 (0.85) [356] | -0.06 (-0.18 to 0.06) | 0.34 |  |  |
|  | ≥75 nmol/L | 1 year | -0.19 (0.84) [187] | -0.06 (0.85) [200] | -0.14 (-0.30 to 0.02) | 0.08 | 0.07 |  |
|  |  | 2 years | 0.05 (0.81) [171] | 0.20 (0.82) [179] | -0.15 (-0.31 to 0.01) | 0.07 |  |  |
|  |  | 3 years | 0.03 (0.76) [196] | 0.14 (0.87) [205] | -0.11 (-0.26 to 0.05) | 0.19 |  |  |

**Abbreviations:** 25(OH)D, 25-hydroxyvitamin D. CI, confidence interval. S.d., standard deviation. n, number.

**Footnotes. 1,** adjusted for baseline value and school of attendance. **2,** median calcium intake 466 mg/day. **3,** deseasonalised values.

### **Table S2.** Mean waist-to-height ratio z-scores at annual follow-up by allocation, main trial participants: overall and by sub-group

|  |  |  | **Vitamin D: mean value (s.d.) [n]** | **Placebo: mean value (s.d.) [n]** | **Adjusted mean difference (95% CI)^1^** | **P for timepoint** | **P for trend** | **P for interaction**  **(treatment*subgroup)** |
| --- | --- | --- | --- | --- | --- | --- | --- | --- |
| Overall |  | 1 year | -0.01 (0.82) [664] | -0.01 (0.81) [659] | -0.02 (-0.10 to 0.07) | 0.67 | 0.01 |  |
|  |  | 2 years | 0.15 (0.83) [610] | 0.19 (0.83) [606] | -0.04 (-0.13 to 0.04) | 0.32 |  |  |
|  |  | 3 years | 0.11 (0.80) [667] | 0.15 (0.83) [686] | -0.05 (-0.14 to 0.03) | 0.22 |  |  |
| By sex | Boys | 1 year | -0.04 (0.76) [304] | -0.12 (0.80) [304] | 0.06 (-0.07 to 0.18) | 0.36 | 0.08 | 0.83 |
|  |  | 2 years | 0.17 (0.76) [284] | 0.12 (0.78) [283] | 0.03 (-0.09 to 0.16) | 0.60 |  |  |
|  |  | 3 years | 0.09 (0.78) [309] | 0.08 (0.78) [325] | -0.01 (-0.13 to 0.11) | 0.85 |  |  |
|  | Girls | 1 year | 0.02 (0.87) [360] | 0.09 (0.80) [355] | -0.09 (-0.21 to 0.03) | 0.14 | 0.08 |  |
|  |  | 2 years | 0.15 (0.88) [326] | 0.26 (0.86) [323] | -0.12 (-0.24 to 0.00) | 0.06 |  |  |
|  |  | 3 years | 0.13 (0.82) [358] | 0.21 (0.87) [361] | -0.10 (-0.21 to 0.02) | 0.11 |  |  |
| By calcium intake^2^ | <median | 1 year | -0.05 (0.81) [314] | -0.09 (0.82) [331] | 0.02 (-0.10 to 0.14) | 0.71 | 0.03 | 0.64 |
|  |  | 2 years | 0.17 (0.79) [288] | 0.13 (0.84) [310] | 0.02 (-0.11 to 0.14) | 0.77 |  |  |
|  |  | 3 years | 0.13 (0.78) [310] | 0.15 (0.80) [358] | -0.02 (-0.14 to 0.10) | 0.74 |  |  |
|  | ≥median | 1 year | 0.01 (0.85) [327] | 0.08 (0.80) [312] | -0.08 (-0.20 to 0.05) | 0.22 | 0.15 |  |
|  |  | 2 years | 0.15 (0.85) [304] | 0.25 (0.82) [279] | -0.10 (-0.23 to 0.03) | 0.12 |  |  |
|  |  | 3 years | 0.09 (0.83) [337] | 0.15 (0.88) [310] | -0.09 (-0.21 to 0.04) | 0.16 |  |  |
| By baseline 25(OH)D concentration^3^ | <75 nmol/L | 1 year | 0.11 (0.82) [349] | 0.04 (0.80) [331] | 0.08 (-0.04 to 0.19) | 0.22 | 0.07 | 0.88 |
|  |  | 2 years | 0.23 (0.84) [323] | 0.21 (0.83) [323] | 0.03 (-0.09 to 0.15) | 0.66 |  |  |
|  |  | 3 years | 0.17 (0.82) [338] | 0.19 (0.80) [356] | -0.02 (-0.14 to 0.10) | 0.78 |  |  |
|  | ≥75 nmol/L | 1 year | -0.17 (0.80) [187] | -0.04 (0.83) [200] | -0.15 (-0.44 to 0.15) | 0.33 | 0.15 |  |
|  |  | 2 years | 0.02 (0.76) [170] | 0.16 (0.77) [179] | -0.15 (-0.46 to 0.16) | 0.34 |  |  |
|  |  | 3 years | 0.01 (0.75) [196] | 0.09 (0.81) [205] | -0.10 (-0.39 to 0.19) | 0.51 |  |  |

**Abbreviations:** 25(OH)D, 25-hydroxyvitamin D. CI, confidence interval. S.d., standard deviation. n, number.

**Footnotes. 1,** adjusted for baseline value and school of attendance. **2,** median calcium intake 466 mg/day. **3,** deseasonalised values.

### **Table S3.** Fat mass and fat-free soft tissue mass at 3-year follow-up by allocation, sub-study participants: overall and by sub-group

|  |  | **Vitamin D: mean value (s.d.) [n]** | **Placebo: mean value (s.d.) [n]** | **Adjusted mean difference (95% CI)^1^** | **P** | **P for interaction** |
| --- | --- | --- | --- | --- | --- | --- |
| **Fat mass, kg** | | | | | | |
| Overall |  | 6.04 (3.86) [202] | 6.28 (3.99) [189] | -0.15 (-0.51 to 0.20) | 0.40 |  |
| By sex | Boys | 4.80 (3.11) [97] | 4.78 (3.05) [89] | 0.18 (-0.30 to 0.66) | 0.45 | 0.14 |
|  | Girls | 7.19 (4.13) [105] | 7.61 (4.27) [100] | -0.45 (-0.96 to 0.05) | 0.08 |  |
| By calcium intake^2^ | <median | 5.73 (3.32) [94] | 5.74 (3.55) [97] | 0.04 (-0.42 to 0.49) | 0.88 | 0.34 |
|  | ≥median | 6.41 (4.35) [102] | 6.88 (4.41) [89] | -0.34 (-0.90 to 0.23) | 0.24 |  |
| By baseline 25(OH)D concentration^3^ | <75 nmol/L | 6.74 (4.28) [103] | 6.64 (4.23) [96] | -0.32 (-0.85 to 0.21) | 0.24 | 0.54 |
|  | ≥75 nmol/L | 5.39 (3.47) [60] | 5.85 (3.71) [54] | -0.04 (-0.62 to 0.55) | 0.91 |  |
| By baseline fat mass | <median | 3.72 (1.08) [103] | 3.65 (1.20) [90] | 0.02 (-0.24 to 0.27) | 0.88 | 0.42 |
|  | ≥median | 8.46 (4.20) [99] | 8.67 (4.15) [99] | -0.27 (-0.91 to 0.37) | 0.41 |  |
| **Fat-free soft tissue mass, kg** | | | | | | |
| Overall |  | 13.22 (2.87) [202] | 13.07 (2.70) [189] | -0.13 (-0.42 to 0.15) | 0.36 |  |
| By sex | Boys | 14.00 (2.87) [97] | 13.54 (3.03) [89] | -0.01 (-0.43 to 0.41) | 0.96 | 0.08 |
|  | Girls | 12.50 (2.69) [105] | 12.64 (2.29) [100] | -0.28 (-0.63 to 0.06) | 0.10 |  |
| By calcium intake^2^ | <median | 13.31 (2.68) [94] | 13.08 (2.67) [97] | -0.09 (-0.48 to 0.30) | 0.67 | 0.68 |
|  | ≥median | 13.23 (3.07) [102] | 13.16 (2.69) [89] | -0.16 (-0.59 to 0.27) | 0.48 |  |
| By baseline 25(OH)D concentration^3^ | <75 nmol/L | 13.33 (3.06) [103] | 13.33 (2.79) [96] | -0.21 (-0.62 to 0.21) | 0.33 | 0.91 |
|  | ≥75 nmol/L | 13.30 (3.03) [60] | 13.01 (2.72) [54] | -0.17 (-0.68 to 0.34) | 0.51 |  |
| By baseline fat-free soft tissue mass | <median | 11.06 (1.54) [89] | 11.40 (1.53) [103] | -0.13 (-0.43 to 0.16) | 0.38 | 0.95 |
|  | ≥median | 14.92 (2.52) [113] | 15.06 (2.43) [86] | -0.14 (-0.60 to 0.33) | 0.57 |  |

**Abbreviations:** 25(OH)D, 25-hydroxyvitamin D. CI, confidence interval. S.d., standard deviation. n, number.

**Footnotes. 1,** adjusted for baseline value and school of attendance. **2,** median calcium intake 466 mg/day. **3,** deseasonalised values.

### **Table S4.** Spirometric outcomes at 3-year follow-up by allocation, sub-study participants: overall and by sub-group

|  |  | **Vitamin D: mean value (s.d.) [n]** | **Placebo: mean value (s.d.) [n]** | **Adjusted mean difference (95% CI)^1^** | **P** | **P for interaction** |
| --- | --- | --- | --- | --- | --- | --- |
| **% predicted FEV1** | | | | | | |
| Overall |  | 81.46 (13.37) [200] | 82.48 (14.54) [187] | -1.68 (-4.23 to 0.88) | 0.20 |  |
| By sex | Boys | 80.33 (11.53) [95] | 83.15 (12.94) [89] | -2.95 (-6.19 to 0.29) | 0.07 | 0.16 |
|  | Girls | 82.48 (14.81) [105] | 81.87 (15.90) [98] | -0.50 (-4.30 to 3.31) | 0.80 |  |
| By calcium intake^2^ | <median | 82.97 (12.53) [92] | 83.31 (15.19) [97] | -1.40 (-4.98 to 2.18) | 0.44 | 0.95 |
|  | ≥median | 80.58 (13.85) [102] | 81.79 (13.90) [87] | -1.44 (-5.21 to 2.33) | 0.45 |  |
| By baseline 25(OH)D concentration^3^ | <75 nmol/L | 82.26 (11.99) [103] | 82.98 (14.21) [95] | -0.34 (-3.72 to 3.05) | 0.84 | 0.10 |
|  | ≥75 nmol/L | 80.10 (12.14) [59] | 84.34 (14.23) [54] | -4.80 (-9.12 to -0.49) | 0.03 |  |
| **% predicted FVC** | | | | | | |
| Overall |  | 86.74 (13.53) [200] | 87.10 (13.51) [187] | 0.23 (-2.30 to 2.76) | 0.86 |  |
| By sex | Boys | 87.77 (12.16) [95] | 90.36 (11.64) [89] | -2.17 (-5.37 to 1.03) | 0.18 | 0.049 |
|  | Girls | 85.81 (14.65) [105] | 84.13 (14.43) [98] | 2.11 (-1.73 to 5.95) | 0.28 |  |
| By calcium intake^2^ | <median | 87.51 (12.47) [92] | 87.15 (13.86) [97] | 0.14 (-3.40 to 3.68) | 0.94 | 0.97 |
|  | ≥median | 86.06 (14.05) [102] | 87.19 (13.33) [87] | 0.62 (-3.08 to 4.33) | 0.74 |  |
| By baseline 25(OH)D concentration^3^ | <75 nmol/L | 86.67 (12.90) [103] | 87.19 (13.84) [95] | -0.22 (-3.73 to 3.30) | 0.90 | 0.92 |
|  | ≥75 nmol/L | 87.08 (12.79) [59] | 86.85 (11.53) [54] | 0.61 (-3.42 to 4.65) | 0.77 |  |
| **% predicted FEV1/FVC** | | | | | | |
| Overall |  | 96.98 (14.02) [200] | 96.99 (10.60) [187] | -0.84 (-3.30 to 1.62) | 0.50 |  |
| By sex | Boys | 97.31 (15.28) [95] | 96.86 (10.03) [89] | 0.02 (-3.64 to 3.69) | 0.99 | 0.67 |
|  | Girls | 96.69 (12.83) [105] | 97.12 (11.14) [98] | -1.32 (-4.58 to 1.94) | 0.43 |  |
| By calcium intake^2^ | <median | 97.64 (11.71) [92] | 97.96 (9.54) [97] | -0.47 (-3.46 to 2.52) | 0.76 | 0.95 |
|  | ≥median | 96.80 (15.07) [102] | 96.12 (11.70) [87] | -1.42 (-5.26 to 2.43) | 0.47 |  |
| By baseline 25(OH)D concentration^3^ | <75 nmol/L | 97.39 (10.36) [103] | 97.39 (10.73) [95] | -0.38 (-3.26 to 2.50) | 0.80 | 0.16 |
|  | ≥75 nmol/L | 95.63 (13.96) [59] | 99.44 (9.48) [54] | -4.47 (-8.82 to -0.12) | 0.04 |  |

**Abbreviations:** 25(OH)D, 25-hydroxyvitamin D. FEV1, forced expiratory volume in 1 second. FVC, forced vital capacity. CI, confidence interval. S.d., standard deviation. n, number.

**Footnotes. 1,** adjusted for baseline value and school of attendance. **2,** median calcium intake 466 mg/day. **3,** deseasonalised values.
